# Exploring the negative triad of childhood maltreatment, fear of relapse, and low sleep quality in multiple sclerosis

**DOI:** 10.64898/2026.08.31.26361813

**Authors:** Nora Trepel, Manuela Gander, Anna Buchheim, Alexander Karabatsiakis

## Abstract

**Background:** Multiple sclerosis (MS) is a chronic, immune-mediated disease of the central nervous system marked by demyelination and neurodegeneration. Beyond physical symptoms, MS is often linked to clinically relevant sleep disturbances. The variability and unpredictability of symptoms and disease progression can also fuel fear of relapse (FoR), undermining well-being and potentially increasing morbidity through inflammatory processes. Understanding biopsychosocial risk factors, including childhood maltreatment (CM) and sleep, in relation to FoR remains an important gap in MS management and research.

**Methods:** Data from *N* = 48 participants were collected via an online survey. We used the *Pittsburgh Sleep Quality Index* (PSQI), the *Fear-of-Relapse Scale* (FoR), and the *Childhood Trauma Questionnaire* (CTQ) to assess the variables of interest. In addition, time points of exposure to different CM subtypes were assessed. Linear regression analyses were conducted to examine associations within the proposed negative triad.

**Results:** A significant negative association between overall sleep quality and FoR was observed. In the total cohort, the interaction between CM and sleep was not a significant predictor of FoR. However, exploratory analysis revealed a significant interaction between CM and sleep among male participants, whereas the same interaction was not significant among female participants.

**Conclusion:** A history of CM and impaired sleep quality introduce new stressors in managing one’s own illness that have received little attention to date. However, the present study found that these factors were at least partly influential on the FoR. The results underscore the translational need for additional support services to enhance prevention and personalized care.

## 1. Introduction

Multiple sclerosis (MS) is one of the most common diseases of the central nervous system (CNS), affecting approximately 2.8 million people worldwide (Multiple Sclerosis International Federation, 2022). Women are affected at least twice as often as men (Gilmour et al., 2018; Multiple Sclerosis International Federation, 2022). The global average age at disease onset is 32 years, with onset ranging from 20 to 50 years across countries (Multiple Sclerosis International Federation, 2022). Although the exact etiology of MS remains unclear, genetic predisposition and environmental and lifestyle factors appear to interact in complex ways, thereby determining an individual’s risk of developing the disease (Goodin, 2014; Olsson et al., 2017).

Although MS occurs more frequently in the families of affected individuals, it follows a multigenic inheritance pattern, and variants in the human leukocyte antigen (HLA) regions are a known risk factor (Hollenbach & Oksenberg, 2015; Lill & Zipp, 2012; Schmidt et al., 2022). In addition to genetic predispositions, several environmental and lifestyle factors contribute to susceptibility to and the course of MS. Risk factors include infection with the Epstein-Barr virus (EBV; Jacobs et al., 2020; Munger et al., 2011), lower vitamin D intake (Balasooriya et al., 2024; Pierrot-Deseilligny & Souberbielle, 2017), and smoking (Hedström et al., 2009; Poorolajal et al., 2017). While the effects of stressful life events (SFLEs), adverse childhood experiences (ACEs), and childhood maltreatment (CM) on MS susceptibility are less clear, many studies suggest significant associations (Eid et al., 2022; Jiang et al., 2020; Nielsen et al., 2014; Rehan et al., 2023).

Common MS symptoms include sensory, visual, and gait disturbances, bowel and urinary dysfunction, sexual dysfunction, dizziness, fine motor skill impairment, sleep disturbances, and fatigue (Egli, 2011; Gelfand, 2014; Schmidt et al., 2022). Additionally, those affected are at higher risk of developing anxiety and depression (Boeschoten et al., 2017; Peres et al., 2022). Because the illness course is heterogeneous and symptoms vary widely, predicting individual disease trajectories remains challenging (Dickerson, 2023). This uncertainty can cause substantial emotional distress among those affected. Although the course of the disease is highly variable, modifiable factors such as smoking (Heydarpour et al., 2018), physical activity (Grosu et al., 2025; Motl & Gosney, 2008; Snook & Motl, 2009), and stress (Drathen et al., 2024; Terrim et al., 2025) are known to influence disease activity and progression.

Given the neuroinflammatory nature of MS (Haase & Linker, 2021) and the strong link between stress-related conditions, such as anxiety and depression, and inflammation (Kim et al., 2022; Renna et al., 2018), the heightened prevalence of psychiatric comorbidities is alarming. Consistent with the increased prevalence of depression, individuals with MS also face a higher risk of suicidality and suicidal ideation than the general population (Jellinger, 2024; Manouchehrinia et al., 2016). Therefore, it is important to expand research efforts to include psychosocial factors, especially because resilience factors such as social support can mitigate the negative effects of stress on relapse risk and improve health-related quality of life and symptoms of anxiety and depression (Brown et al., 2022; Costa et al., 2012; Henry et al., 2019; Ratajska et al., 2020). Identifying individuals for whom coping with the disease is particularly challenging and for whom this uncertainty causes significant anxiety represents a significant research gap and a key aspect of predictive, preventive, and personalized medicine (3PM). Although research on MS pathophysiology and treatment has advanced considerably (Dighriri et al., 2023), patients still report a significantly lower quality of life than the general population (Amtmann et al., 2018; Li et al., 2022). To treat MS holistically, research must encompass the full range of factors that shape the lives of those affected. A deeper understanding of these factors can pave the way for more accurate prediction of which individuals are most severely affected. This also enables preventive approaches by allowing more precise early assessment and treatment of an individual’s distress, potentially preventing secondary symptoms. Additionally, a more comprehensive understanding enables more personalized treatment through targeted interventions that address individual stressors and needs. To support a more bio-psycho-socially oriented approach to care, it appears necessary to examine not only biological but also psychosocial functioning in those affected. This includes coping with uncertainty about the disease’s course (Alschuler & Beier, 2015; Dickerson, 2023). Specifically, the so-called fear of relapse (FoR) is a significant source of stress for individuals affected by MS (Khatibi et al., 2020, 2021). The FoR encompasses cognitive, affective, and behavioral components, including beliefs about the likelihood of progressive disability or worsening symptoms, the associated emotional distress, and resulting avoidance behaviors (Khatibi et al., 2020).

Various studies have linked FoR or similar constructs to adverse mental health outcomes, including depression, stress, anxiety, fatigue, and reduced quality of life (Khanlari Aziz et al., 2023; J. Nielsen et al., 2018, 2022). Although several studies have reported significant associations between FoR and reduced quality of life (Khatibi et al., 2021; Mallahzadeh et al., 2025; Öztürk & Öztürk, 2026), the full integration of FoR into a bio-psycho-social framework and the inclusion of risk factors such as sleep (quality and duration) and CM remain underexplored. Understanding the causes, correlates, and consequences of FoR enables the development of targeted, personalized interventions to prevent the chronification of stress and related disorders, such as anxiety and depression, and to provide care that accounts for their inflammatory effects.

In general, individuals with MS experience poorer sleep quality across multiple domains than those without MS (Moradi et al., 2025). More than half of individuals with MS report impaired sleep quality (Alis et al., 2025; Yun et al., 2025). Several studies report associations between sleep quality and mental and physical health, cognitive impairments, and overall quality of life in individuals with MS (Golabi et al., 2024; Kotterba et al., 2018; Laslett et al., 2022; Lee et al., 2021; Sarraf et al., 2014). In addition, lower sleep quality is associated with higher anxiety levels (Alis et al., 2025; Ozdogar et al., 2025), which is especially important given the heightened health anxiety among those affected (Alberts et al., 2011; Hayter et al., 2016). Notable associations have also been demonstrated between resilience and sleep quality (Novak & Lev-Ari, 2023). Poor sleep also appears to affect the social networks of persons with MS and their experience of social support (Riegler et al., 2024), underscoring its crucial and extensive role. However, the reported direction of the relationships between sleep and mental health varies. For example, a longitudinal study of the general population confirmed the negative impact of insomnia on anxiety and depression (Johansson et al., 2021). Furthermore, there is evidence of a bidirectional relationship in which sleep disturbances and anxiety mutually influence or reinforce each other (Peng et al., 2024). Given the neuroinflammatory nature of MS, it is important to note that multiple domains of poor sleep quality are also linked to systemic inflammation (Engert & Besedovsky, 2025; Irwin et al., 2016; Jin et al., 2023; Petrov et al., 2020).

Sleep disturbances can be influenced by many factors, including childhood adversity (Simon & Admon, 2023) and stressful life events (Kalmbach et al., 2018; Lo Martire et al., 2020). The strong association between childhood adversity, particularly CM, and impaired sleep quality has been repeatedly demonstrated in the general population (Brown et al., 2022; Schønning et al., 2022) and, although less extensively studied, in individuals diagnosed with MS (Goldman et al., 2025). Research indicates that sleep disturbances associated with child maltreatment are evident in children and adolescents, who show increased odds of insomnia symptoms, reduced sleep duration, and more frequent nightmares (Schønning et al., 2022). A key mechanism linking childhood adversity and sleep is dysregulation of the hypothalamic-pituitary-adrenal (HPA) axis (Simon & Admon, 2023). Simon and Admon (2023) conclude that sleep disturbances and HPA axis dysfunction reinforce each other, thereby increasing stress vulnerability. Accordingly, the relationship between stress, which is closely related to HPA axis dysfunction, and sleep appears to be bidirectional and mutually reinforcing (Lo Martire et al., 2020; Simon & Admon, 2023; Yap et al., 2020).

For those affected by MS, poor sleep quality exacerbates distressing factors such as anxiety, depression, and impaired cognitive function. It also weakens protective factors, such as resilience, that support coping with the disease. Poor sleep quality predicts subsequent cognitive decline (Carpi et al., 2024), cancer (Song et al., 2021), and all-cause mortality risk among middle-aged and older adults (Del Brutto et al., 2024). These findings, especially those related to heightened inflammation, underscore the importance of accounting for subjective sleep quality and childhood adversity in a comprehensive, holistic MS treatment approach that prioritizes the individual’s well-being. Psychosocial care is a common recommendation in various MS treatment and management guidelines (Hemmer et al., 2024; National Institute for Health and Care Excellence, 2022). Accordingly, considering sleep, childhood experiences, and disease-related fears together is central to ensuring such comprehensive care. The relationships described above suggest a close link among sleep, FoR, and childhood adversity. It can therefore be hypothesized that poor-quality sleep exacerbates the FoR and that CM moderates this relationship.

## 2. Methods

Data were collected through announcements on social media, support groups, and flyers distributed at University Hospitals Ulm and Augsburg in Germany, as well as at several local neurology practices in the Innsbruck area in Tyrol (Austria). We conducted the survey online via *LimeSurvey*, and it took approximately 30 minutes to complete. Inclusion criteria were age 18 or older and an MS diagnosis of at least three months at the time of participation. To recruit participants as broadly as possible, we applied no additional formal exclusion criteria. Before the survey questions began, participants were informed about the procedure, benefits, and potential burdens. At any time, participants could quit or pause the questionnaire. Participants could create a unique pseudonymization code to resume the survey later. Data collection was conducted anonymously. The local Ethics Review Board at Innsbruck University approved the study protocol.

### 2.1 Measures

The *Fear-of-Relapse Scale* by Kathibi et al. (2020) was used to assess FoR. It is a self-report instrument with 26 items. In addition to anxious thoughts about relapse and its consequences, the items assess avoidance behaviors. Items are rated on a five-point *Likert* scale. In the validation study, internal consistency was very high (*α* = .92), and test-retest reliability (*r* = .74) indicated the scale’s relative stability. The English version was translated into German, and the German version showed excellent internal consistency in this study (*α* = .91).

Sleep quality was assessed using the *Pittsburgh Sleep Quality Index* (PSQI) self-report questionnaire, originally developed in English by Buysse et al. (1989). The PSQI demonstrates high specificity and sensitivity, with high internal consistency (*α* = .83) and high test-retest reliability (*r* = .85 to .87; Backhaus et al., 2002; Buysse et al., 1989). In this study, the German version by Riemann and Backhaus (1996) was used. The PSQI comprises 19 items grouped into seven components, which are summed to produce an overall score. The components include *Subjective Sleep Quality*, *Sleep Latency*, *Sleep Duration*, *Habitual Sleep Efficiency*, *Sleep Disturbances*, *Use of Sleeping Medication*, and *Daytime Dysfunction*. For both the individual components and the overall score, higher scores indicate poorer sleep quality. Internal consistency in this study was acceptable (*α* = .67).

The extent and distribution of CM were assessed using the *Childhood Trauma Questionnaire* (CTQ) by Bernstein et al. (1994), in its established short form (CTQ-SF), which comprises 28 items (Bernstein et al., 2003; Bernstein & Fink, 1998). This self-report questionnaire is considered a valid screening tool for assessing experiences of abuse and neglect among adolescents and adults aged 12 and older. Specifically, it measures five subscales: *Emotional Abuse*, *Physical Abuse*, *Sexual Abuse*, *Emotional Neglect*, and *Physical Neglect*. By summing the items within each subscale and across all subscales, total scores can be calculated and interpreted as quantitative measures of CM severity. According to Bernstein and Fink (1998), severity-level classifications can also be made. Based on this classification, individuals can be categorized as having experienced CM (CM^+^). With a mild cutoff, individuals can be considered CM^+^ if they obtain a low-to-moderate total score on at least one subscale. With a moderate cutoff, a person is considered CM^+^ only if they obtain a moderate-to-severe total score on at least one subscale. For each subscale, the described severity classification was applied, followed by classification as CM^+^ or CM^-^. This also determined whether a subscale was considered fulfilled. The *Maltreatment Load* is calculated by summing the scores on the fulfilled subscales.

The German version of the CTQ was validated by Klinitzke et al. (2012) in a representative sample of the general population (*N* = 2,500). The 4-factor structure showed adequate model fit, and the subscales (except *Physical Neglect*) exhibited high internal consistency (*α* ≥ .80). Construct validity was further supported by negative correlations with quality of life and positive correlations with anxiety and depression (Klinitzke et al., 2012). As in the validation study, *Physical Neglect* showed the lowest internal consistency (*α* = .70), whereas the other subscales and the total score had good to excellent internal consistency (*α* > .86). To assess CM in greater detail, the CTQ response format was combined with a time scale, as provided in the *Maltreatment and Abuse Chronology of Exposure* (MACE). Respondents are asked to check all years in which the event described in the respective item occurred. This situates the experience in a temporal context and specifies the duration of CM exposure (Teicher & Parigger, 2015).

### 2.2 Participants and statistical analysis

A total of *N* = 53 individuals participated in this study. Because the CTQ includes questions about potentially traumatic childhood experiences, participants could skip items and continue the survey without providing additional information. Participants who answered fewer than four items per subscale were excluded from the analysis. This applied to *n* = 5 participants; accordingly, the analyses were conducted with *n* = 48. If only one item per person was missing in a subscale, Little’s MCAR test (*Missing Completely at Random*) was used to assess whether the missing values were randomly distributed (Little, 1988). This applied to 8 items in total. The results of Little’s MCAR test were not significant for any of the subscales, suggesting that the missing values can be assumed to be randomly distributed. Based on this assumption, the missing values were replaced using mean imputation and were manually checked for plausibility.

The data were analyzed using version 4.5.1 of the statistical software R (R Core Team, 2025). In accordance with the requirements of the specific tests, we checked the data for normality, homogeneity of variance, multicollinearity, and heteroscedasticity before analysis. Normality was assessed using the Shapiro-Wilk test, with histograms and Q-Q plots for graphical inspection. Homogeneity of variance was tested before the t-tests using *Levene’s* test. Multicollinearity was assessed using *variance inflation factors* (VIF). Homoscedasticity was assessed in the regression analysis using a residual-vs.-fitted plot. If these conditions were not met, appropriate nonparametric tests were selected. The significance level was set at *α* = 0.05, although results slightly above this level are also considered relevant if the effect size is sufficiently large (Lee, 2016; Sullivan & Feinn, 2012). The hypothesis was tested using a linear regression model, with the total PSQI score as the predictor and FoR as the outcome variable. In addition, a moderation analysis was conducted in which FoR served as the predictor, and the interaction between the total PSQI and CTQ scores was examined. As part of an exploratory analysis to generate hypotheses, this moderation was also analyzed separately for men and women.

### 2.4 Sociodemographics and characterization of the cohort

Participants’ ages ranged from 19 to 73 years (*M* = 43.54, *SD* = 13.54). Most participants identified as female (79%). The disease course was predominantly relapsing-remitting (69%), and medication was usually taken continuously (69%). Total disease duration ranged from 3 to 480 months (*M* = 103.33, *SD* = 109.19). Three outliers were removed due to implausibility (*n* = 45). Over half of the sample (73%) reported engaging in physical activity three to four times per week, and only 10 % reported smoking tobacco daily. The average BMI was *M* = 25.7 (*SD* = 4.5). More detailed sociodemographic information is provided in Table 1.

**Table 1:** Sociodemographic data of the study cohort.

| Variable | Extent |
| --- | --- |
| Sex (% ($n$)) | Female 79% (38), male 21% (10) |
| Age (years) | $M = 43.54$ ($SD = 13.61$), range: 19-73 |
| Disease form (% ($n$)) | RRMS: 69% (33), PRMS: 13% (6), SPMS: 15% (7), unknown: 3% (2) |
| Continuous medication (% ($n$)) | 69% (33) |
| Disease duration in months ($n = 45$) | $M = 103.33$ ($SD = 109.19$), range: 3 - 480 |
| Physical activity $\geq 3$x/week (% ($n$)) | 73% (35) |
| BMI ($\text{kg}/\text{m}^2$) | $M = 25.71$ ($SD = 4.5$), range: 18.47 – 36.92 |
| Daily smoking (% ($n$)) | 10% (5) |

*Note.* $M$ = Mean, $SD$ = Standard deviation, RRMS = Relapsing-remitting multiple sclerosis, PRMS = Primary progressive multiple sclerosis, SPMS = Secondary progressive multiple sclerosis.

### 2.5 Adverse childhood experiences and maltreatment load

Figure 1 shows the prevalence of different forms of CM and the percentage distribution of CM across childhood and adolescence. *Emotional Neglect* (*M* = 11.41, *SD* = 6.09) and *Emotional Abuse* (*M* = 10.37, *SD* = 5.44) were the most prevalent forms of CM. On average, CM occurred between 10.27 and 12.36 years of age. For each subscale, the severity classification described above was applied. Using both mild and moderate cutoffs, more than half of the sample was classified as CM^+^ (71%, 52%, respectively).

**Figure 1:**
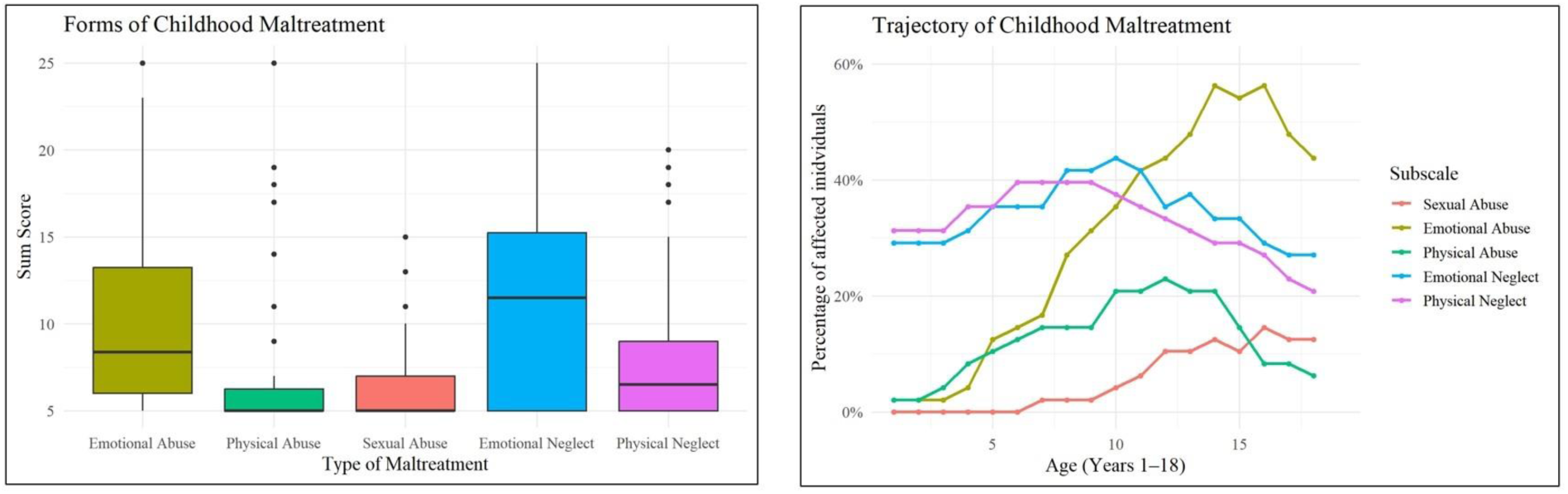
Prevalence of different forms of CM in the overall sample and over time.

Overall, participants met criteria for an average of *M* = 1.88 (*SD* = 1.61) subscales (*M* = 1.08; *SD* = 1.32 with a moderate cut-off). A more detailed description of the distribution of the *Maltreatment Load*, as well as a comparison between the mild and moderate cut-offs, is presented in Figure 2.

**Figure 2:**
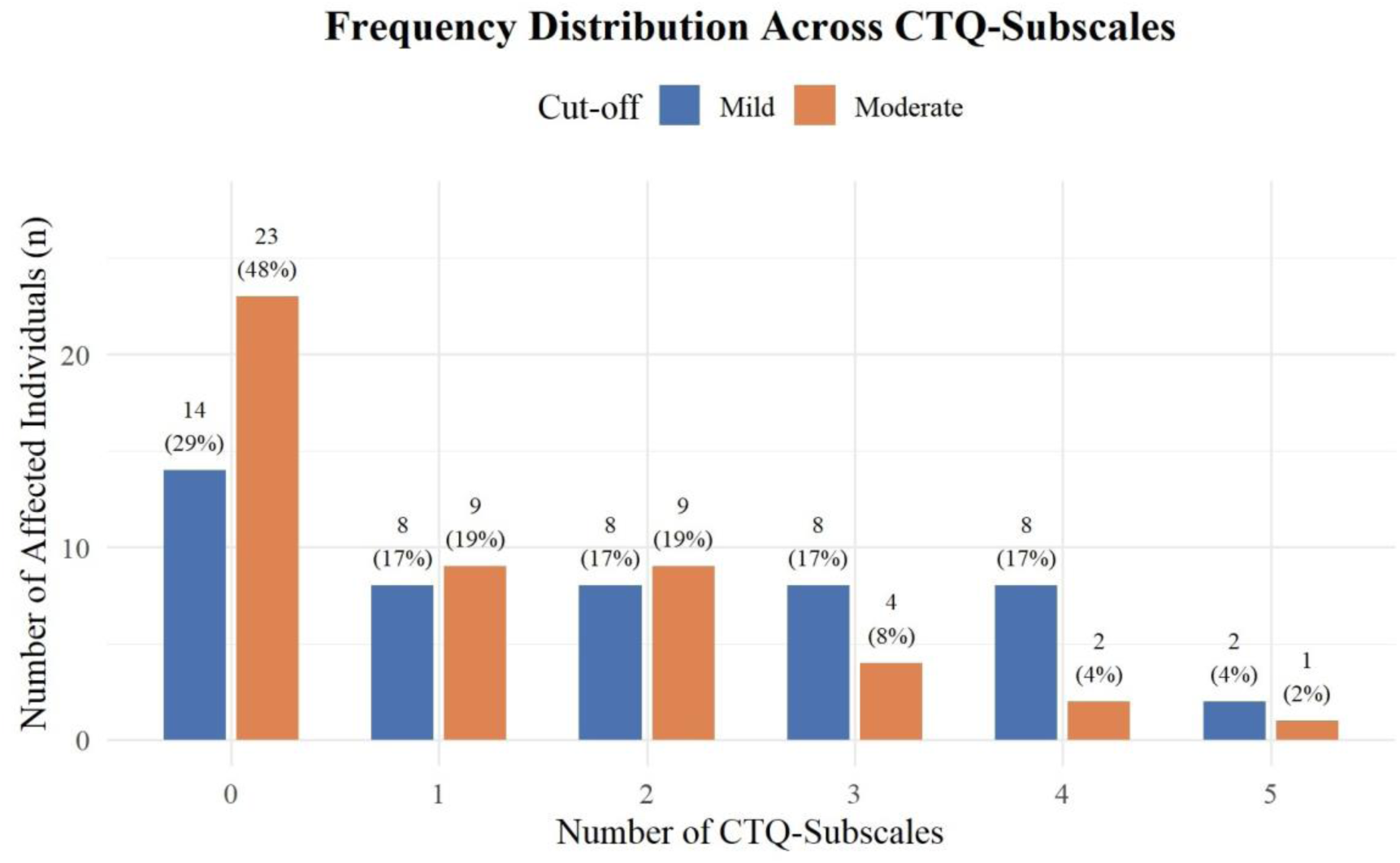
Frequency distribution across CTQ-Subscales.

### 2.6 Fear of relapse and sleep quality

The mean FoR score for the entire sample was 34.35 (*SD* = 16.14). Individuals with different disease courses did not differ significantly in their fear of new relapses, *H*(48) = 5.43, *df* = 3, *p* = .143, *η*2 = 0.055. In the overall sample, 58% of participants scored above the cutoff of > 5 for good sleep quality. The subscales *Daytime Sleepiness* (*M* = 1.69, *SD* = 0.78), *Sleep Latency* (*M* = 1.33, *SD* = 1.06), and *Sleep Disturbances* (*M* = 1.25, *SD* = 0.53) were the most pronounced (Table 2). Using a moderate cutoff, subjects with a history of CM (CM^+^) reported significantly higher *Daytime Dysfunction* and overall scores (Table 2).

**Table 2:** Sleep quality scores (PSQI) for the overall sample of subjects with multiple sclerosis and for group differences between those with a history of CM (CM^+^) and those without CM (CM^-^). Higher values indicate poorer sleep quality.

| Scale<br>(range) | Total<br>M (SD) | CM<sup>-</sup><br>M (SD) | CM<sup>+</sup><br>M (SD) | Group Differences |
| --- | --- | --- | --- | --- |
| *Subjective Sleep Quality* (0-3) | 1.06<br>(0.63) | 1.00<br>(0.60) | 1.12<br>(0.66) | $W = 259, Z = -0.67, p = .501, r = -0.10$ |
| *Sleep Latency* (0-3) | 1.33<br>(1.06) | 1.74<br>(1.11) | 1.48<br>(1.00) | $W = 237.5, Z = -1.07, p = .285, r = -0.15$ |
| *Sleep Duration* (0-3) | 0.38<br>(0.67) | 0.39<br>(0.58) | 0.36<br>(0.76) | $W = 313, Z = -0.66, p = .608, r = -0.07$ |
| *Habitual Sleep Efficiency* (0-3) | 0.75<br>(1.08) | 0.65<br>(0.98) | 0.84<br>(1.18) | $W = 266, Z = -0.51, p = .608, r = -0.07$ |
| *Sleep Disturbances* (0-3) | 1.25<br>(0.53) | 1.13<br>(0.46) | 1.36<br>(0.57) | $W = 235, Z = -1.40, p = .162, r = -0.20$ |
| *Use of Sleeping Medication* (0-3) | 0.48<br>(1.07) | 0.39<br>(1.03) | 0.56<br>(1.12) | $W = 259, Z = -0.87, p = .386, r = -0.013$ |
| *Daytime Dysfunction* (0-3) | 1.69<br>(0.78) | 1.39<br>(0.66) | 1.96<br>(0.79) | $W = 168, Z = -2.66, \mathbf{p = .008}, r = -0.38$ |
| Overall score (0 – 21) | 6.94<br>(3.48) | 6.74<br>(4.23) | 8.52<br>(3.95) | $W = 207.5, Z = -1.66, p = .097, r = -0.24$ |

Note. *M* = Mean, *SD* = Standard deviation. Results in bold are significant. The classification CM<sup>-/+</sup> is based on the moderate cutoff.

## 3. Results

First, a simple linear regression tested whether PSQI scores significantly predicted FoR scores. The regression was statistically significant, with *R^2^* = 0.17, *adj. R^2^* = 0.16, *F*(1, 46) = 9.73, *p* = .003, confirming that PSQI scores significantly predicted FoR scores (*b* = 6.75, *SE* = 2.16, *t*(46) = 3.12, *p* = .003). Accordingly, a one-SD increase in PSQI scores was associated with a 6.75-point increase in FoR. The Q-Q plot indicated that the residuals were approximately normally distributed, and the residuals vs. fitted plot showed no substantial violations of homoscedasticity.

Next, we used multiple linear regression to test whether CTQ scores moderated the association between PSQI and FoR scores. The overall model accounted for 16% of the variance in FoR scores, with *R^2^* = 0.21 and *adj. R^2^* = 0.16, *F*(3, 44) = 3.96, *p* = .014, after adjusting for model complexity. Childhood maltreatment did not moderate the association between sleep and FoR (*b* = 0.60, SE = 2.27, *t*(44) = 0.27, *p* = .792). The main effect of PSQI scores was significant (*b* = 6.01, *SE* = 2.23, *t*(44) = 2.70, *p* = .010), whereas the main effect of CTQ scores was not (*b* = 3.08, *SE* = 2.26, *t*(44) = 1.37, *p* = .179). The Q-Q plot showed no strong deviations from normality. All predictors in the regression model had VIFs close to 1 (PSQI = 1.06, CTQ = 1.09, PSQI*CTQ = 1.03), indicating low multicollinearity, and the residuals- vs.-fitted plot revealed no substantial violations of homoscedasticity. Therefore, the data met the statistical prerequisites.

As an exploratory analysis, moderation was assessed separately for men and women to determine whether relevant effects emerged. The overall model, using only male participants (*n* = 10), accounted for 68% of the variance in FoR scores (R^2^ = 0.79, *adj. R^2^* = 0.68, *F*(3, 6) = 7.36, *p* = .020). The interaction between PSQI and CTQ scores was significant (*b* = 25.67, *SE* = 8.19, *t*(6) = 3.13, *p* = .020), suggesting that CM moderates the relationship between sleep quality and FoR. While the main effect of PSQI scores remained significant in this model (*b* = 17.25, *SE* = 5.00, *t*(6) = 3.45, *p* = .014), the main effect of CTQ scores was not significant (*b* = 0.11, *SE* = 4.91, *t*(6) = 0.02, *p* = .983). The Q-Q plot showed slight tail deviation, but the Shapiro-Wilk normality test was not significant (*W* = 0.95, *p* = .702). Therefore, an approximately normal distribution can be expected. All predictors in the regression model had VIFs below 2 (PSQI = 1.45, CTQ = 1.39, PSQI*CTQ = 1.05), indicating low multicollinearity. The residuals- vs-fitted plot suggested possible violations of homoscedasticity.

The overall model for women (*n* = 38) was not significant (*R^2^* = 0.16, *adj. R^2^* = .08, *F*(3, 34) = 2.09, *p* = .119). The interaction between PSQI and CTQ scores in this model was also not significant (*b* = -0.86, *SE* = 2.21, *t*(34) = -0.39, *p* = .699). Although PSQI scores did not significantly predict FoR scores, they showed a trend (*b* = 4.28, *SE* = 2.32, *t*(34) = 1.84, *p* = .074). As in the previous models, the main effect of CTQ scores was not significant (*b* = 3.28, *SE* = 2.37, *t*(34) = 1.39, *p* = .175). The Q-Q plot showed no strong deviations from normality, and the Shapiro-Wilk normality test was not significant (*W* = 0.96, *p* = .254). All predictors in the regression model had VIFs close to 1 (PSQI = 1.03, CTQ = 1.07, PSQI*CTQ = 1.04), indicating low multicollinearity, and the residuals-vs.-fitted plot revealed no substantial violations of homoscedasticity.

## 4. Discussion

This study examined how sleep quality affects fear of relapse and whether this relationship is moderated by childhood maltreatment. The aim was to deepen understanding of how CM and impaired sleep quality affect individuals diagnosed with MS, to further elaborate on the concept of fear of relapse, and to identify potential influential factors. Sleep quality showed a significant association with fear of new relapses, while the effect of CM appeared to vary. Among male participants, however, CM moderated the relationship between sleep quality and FoR. Interestingly, CM did not predict FoR *per se*. Impaired sleep quality is highly prevalent among individuals with MS (Moradi et al., 2025) and may influence disease course (Buratti et al., 2019; Sahraian et al., 2017). Numerous heterogeneous studies in recent years have examined the impact of childhood adversity on disease course and clinical outcomes (Drathen et al., 2024; Eid et al., 2022; Goldman et al., 2025). Despite the importance of both sleep and childhood adversity, their association has rarely been examined in the context of MS. Uncertainty about disease course places considerable strain on individuals with MS, often resulting in significant fear of new relapses. Therefore, identifying potential risk factors for FoR, such as sleep quality, CM, and their interaction, is crucial to personalized care.

The observed link between reduced sleep quality and increased FoR is consistent with other studies demonstrating the influence of sleep quality on anxiety and depression (Ozdogar et al., 2025; Siengsukon et al., 2018), as well as on the fear of falling in individuals with MS (Abou et al., 2024). However, similar effects appear to operate in the opposite direction. The predictive relevance of psychosocial resilience – and, more specifically, the fear of disease progression – for sleep has also been documented in other disease conditions, including chronic heart failure (Xiong et al., 2023) and endometriosis (Pickup et al., 2024). It therefore stands to reason that the relationship between FoR and sleep quality is mutually reinforcing and closely linked to anxiety and depression. Such a bidirectional relationship is frequently discussed both broadly in the context of sleep and mental health and more specifically in relation to sleep and anxiety or depression (van Dyk et al., 2016; Yasugaki et al., 2025). Therefore, it is necessary to examine the extent to which interventions aimed at improving sleep quality also affect FoR, and vice versa. This also underscores sleep quality as an essential factor in mental health and the importance of including it in personalized, holistic care. Given the discrepancy between objective and subjective sleep quality demonstrated in various studies (Cudney et al., 2022; Fabbri et al., 2021; Pierson-Bartel & Ujma, 2024), future research would benefit from supplementing retrospective self-report measures with ecological momentary assessments (EMAs) or objective measures such as actigraphy. Another important aspect of the close relationship among sleep, CM, and MS is inflammation (Irwin, 2019; Irwin et al., 2016; Kerr et al., 2021). Increased FoR has also been linked to higher stress (Khatibi et al., 2020; Mallahzadeh et al., 2025; Shaygannejad et al., 2021), which is associated with inflammation (Kim et al., 2022; Marsland et al., 2017). Heightened inflammation is particularly problematic in conditions such as MS because, especially in chronic cases, it reduces overall functional ability and impairs quality of life (Gil-González et al., 2020; Guzel et al., 2016; Saçmacı et al., 2021). This underscores the importance of addressing sleep and anxiety in clinical settings to counteract disease progression and deterioration and to build greater resilience.

The absence of a significant interaction between sleep and CM on FoR in this study may reflect the nature of the relationships involved. Most studies suggest that sleep quality mediates rather than moderates the relationship between CM and multiple mental health outcomes (Hillebrant-Openshaw & Wong, 2024; Liu et al., 2023). For example, Laskemoen et al. (2021) reported significantly more frequent insomnia among individuals with childhood trauma and found that insomnia symptoms partially mediated the relationship between childhood trauma and the severity of depressive and anxiety symptoms. Luo et al. (2022) also found a significantly higher prevalence of sleep disorders, specifically insomnia, among individuals who experienced childhood trauma, and a partially mediating effect of insomnia on the relationship between childhood trauma and depression severity. Although few studies have examined the potential interaction between sleep quality and CM, the effects remain unclear.

However, Masuya et al. (2024) found that the interaction between sleep disturbances and CM significantly affected depressive symptoms. Therefore, the absence of an interaction between CM and sleep quality could also reflect a small effect size that may not be detectable in a sample as small as this study’s. The present study, along with the reviewed literature, highlights the interconnectedness between CM and transdiagnostic symptoms such as anxiety and sleep disturbances. These associations and their shared inflammatory mechanisms underscore the need for enhanced bio-psycho-social approaches in MS care.

Our finding that CM moderated the relationship between sleep quality and FoR in men highlights the potential relevance of trauma-related processes for relapse-related fears and raises questions about sex-related differences in coping with the disease. Given documented sex differences in neuropsychiatric symptoms of MS, such as anxiety, depression, and fatigue (Brasanac et al., 2025; Freedman et al., 2026), future research should be sensitive to sex-specific findings.

The role of childhood maltreatment has received little attention in MS care, and trauma-focused treatment approaches are not routinely considered despite the high prevalence of maltreatment experiences in our sample. Our results underscore the importance of a sex-sensitive approach to MS care and suggest that trauma-informed assessment and intervention strategies may be particularly relevant for men with MS. Importantly, several evidence-based psychotherapeutic approaches are available to target trauma-related symptoms and the long-term sequelae of adverse childhood experiences (Rothbaum & Watkins, 2025). Incorporating trauma-informed treatment elements into MS care may therefore represent a novel strategy to reduce FoR and enhance psychological well-being and quality of life, particularly among men with a history of CM.

Pediatric-onset MS, characterized by higher inflammatory disease activity and a more active disease course than adult-onset MS (Kornbluh & Kahn, 2023), may be a particularly important context for trauma-informed care. Given that CM in our sample occurred predominantly in late childhood and early adolescence, an important question for future research is whether trauma exposure during these sensitive developmental periods contributes to psychological vulnerability and disease adaptation in individuals with early-onset MS. If so, trauma-informed interventions may be a particularly valuable, yet underexplored, component of comprehensive MS care, with the potential to improve long-term psychosocial adjustment and quality of life.

In our view, the present findings suggest that fear of relapse in MS should not be conceptualized solely as a rational response to an unpredictable chronic illness, but rather as a complex biopsychosocial phenomenon shaped by interactions among stress regulation, sleep quality, early life adversity, and psychological adaptation to the disease. Consequently, psychotherapeutic interventions should extend beyond symptom management and address the broader mechanisms that contribute to persistent fear and emotional vulnerability. Future research should also examine whether disease-specific therapist competencies contribute to treatment outcomes in patients with elevated FoR. Based on the present findings, several competencies appear particularly relevant.

First, psychotherapists should possess a fundamental understanding of MS, including disease progression, relapse characteristics, fatigue, cognitive symptoms, disease-modifying therapies, and the interactions between stress, sleep, and immune functioning. Such knowledge enables therapists to contextualize illness-related fears realistically and avoid reinforcing maladaptive illness beliefs (Bassi et al., 2020; Scandiffio et al., 2025). Second, therapists should be competent in trauma-informed care, including the recognition of CM, the implementation of stabilization strategies, and the appropriate integration of trauma-focused interventions when clinically indicated. Third, attachment-informed therapeutic skills are likely to be particularly valuable. The ability to establish a secure therapeutic alliance characterized by empathy, emotional availability, consistency, validation, and containment may foster resilience and reduce attachment-related distress activated by disease uncertainty (Ciechanowski et al., 2002). Fourth, therapists should be skilled in promoting mentalization by helping patients reflect on bodily experiences, emotional responses, and cognitive interpretations rather than reacting automatically to perceived physical threats. Finally, given the observed role of sleep quality, psychotherapists should routinely assess sleep disturbances, possess basic knowledge of evidence-based insomnia treatments (Bacaro et al., 2021; Turkowitch et al., 2024; Siengsukon et al., 2026) and collaborate with sleep medicine specialists when appropriate.

Within the context of an unpredictable chronic disease such as MS, disturbed sleep may further amplify physiological and emotional stress responses, thereby contributing to persistent fear of relapse. Psychotherapeutic interventions should therefore extend beyond illness-related anxiety and address sleep regulation, trauma-related vulnerabilities, attachment processes, emotion regulation, and tolerance of uncertainty. Future intervention studies should evaluate whether integrative psychotherapeutic approaches targeting these mechanisms reduce FoR and improve psychological well-being, quality of life, and potentially even inflammatory processes in individuals living with MS.

However, this study has notable limitations. Above all, the sample size is relatively small, especially when including only the male participants. Therefore, the sex-specific results in particular should be interpreted with caution and further investigated in future studies. Additional limitations are the cross-sectional design and self-report data as the sole source of information. Furthermore, the person-based mean imputation used to replace missing values may artificially reduce variance. Although only a few values were missing and they were MCAR, the effects of this variance reduction on the results cannot be ruled out. In addition, the FoR scale was validated only in individuals with a relapsing-remitting disease course but was applied to a mixed sample in this study. The scale demonstrated excellent internal consistency, and individuals with different disease courses did not differ systematically in their FoR. Nevertheless, it remains unclear whether the concept applies to other disease courses or requires a broader scope. Furthermore, validation of a German version of the FoR scale is still pending. Because the variables examined may constitute a complex construct with multiple interplays and possible combinations of moderating and mediating relationships, a larger, more comprehensive sample would be advantageous for testing these relationships. This study provides the first empirically based evidence for the need to better monitor health and function in individuals with MS by addressing bio-psycho-social factors relevant to sleep quality, anxiety, and inflammation. Given that depression and other psychiatric comorbidities are widespread among patients with MS (Boeschoten et al., 2017; Maric et al., 2021), it would be of interest to examine them in the context of adverse childhood experiences and FoR. A possible confounding variable could be the use of psychotherapy or other psychosocial interventions, which could reduce the FoR and also correlate with psychiatric comorbidities. Building on the present findings, future research should examine whether integrative psychotherapeutic interventions targeting sleep quality, trauma-related vulnerabilities, emotion regulation, and tolerance of uncertainty can reduce fear of relapse. Furthermore, studies should investigate the extent to which disease-specific therapist competencies contribute to treatment effectiveness and personalized care in individuals with MS. Accordingly, future studies on FoR should examine the effects of psychotherapy and the role of psychiatric comorbidity and multimorbidity.

We expect strong demand for improved monitoring functions in clinical settings, covering lifestyle factors, cognitive and affective functioning, and variables important for somatic health, including sleep and inflammation, to provide not only a broader but also a more detailed picture of the needs and demands of individuals with MS. A deeper, more nuanced understanding of the burdens inherent in the disease, such as FoR, is essential to provide comprehensive care for those affected. Furthermore, similar correlations between childhood adversity, sleep quality, and concerns regarding disease progression can likely be found in individuals with other chronic diseases. Therefore, these areas require more research.

## 5. Conclusion

This study specifically examined the relationship between sleep and FoR, as well as the role of CM in bridging the research gap regarding the nature and development of FoR and the comprehensive effects of impaired sleep and CM. Enhanced psychosocial care for individuals with MS is a common conclusion of the respective research (Marrie et al., 2015; Morris-Bankole & Ho, 2023) and a recommendation of the Sk2-guideline for the treatment of MS issues by the *German Society of Neurology* (Hemmer et al., 2024) as well as the *NICE guideline for the management of MS* (National Institute for Health and Care Excellence, 2022). However, a clinically relevant deficiency remains in effective, comprehensive psychosocial treatment (Grech et al., 2021), although there are signs of a fundamental improvement in this care (Raissi et al., 2015). Because effective treatment requires accounting for individually varying bio-psycho-social factors, this study makes a partial contribution by not only emphasizing FoR as a relevant concept but also by offering approaches to identify potential risk groups, including poor sleep quality as a predisposing marker for FoR.

## Declarations

### Ethics approval

All procedures were approved by the Review Board for Issues of Ethics in Scientific Research (Certificate of good standing, #45/2025) at the Department of Psychology, University of Innsbruck, Austria.

### Consent to participate

Participants provided informed consent to participate in the online survey.

### Consent for publication

After reviewing the manuscript, all authors approved its final version.

### Funding

This research received no specific grant from funding agencies in the public, commercial, or not-for-profit sectors.

### Competing interests

The authors have no relevant financial or non-financial interests to disclose.

### Data Availability

The datasets generated and analyzed during the current study are available from the corresponding author on reasonable request.

## Notes

### Competing Interest Statement

The authors have declared no competing interest.

